# Prevalence, Patterns and Associated Factors for Neural Tube Defect Among Young Infants Admitted at Bugando Medical Centre in Northwestern Tanzania: A Cross Section Hospital Based Observational Study

**DOI:** 10.1101/2025.06.19.25329971

**Authors:** Caster Gigwa Justine, Rose Laisser, Florentina Mashuda, Ally Tuwa, Cosmas Clement Nestory, Adolphine Hokororo, Kija Malale, Benison R kidenya, Enock Diocle, Furaha Asaeli Mwampiki, Hyasinta Jaka, John Kaswija, Asnath Mpelo, Haruna Dika

**Author notes:** Corresponding Author: Caster Gigwa Justine.

## Abstract

**Background:** In Africa, including Tanzania, the trend for neural tube defects (NTDs) has been increasing, and has been shown to increase morbidity and mortality in young infants.

At Bugando Medical Centre (BMC)-a tertiary hospital serving the population of Northwestern Tanzania, the cases of NTD are commonly reported, but its exact prevalence, patterns and associated factors in young infants admitted at BMC remain unknown. Therefore, this study determined the prevalence, patterns and associated factors for NTD among young infants admitted at BMC.

**Methodology:** It is a cross-sectional observational study involving young infants aged 0-3 months admitted at BMC. Ethical approval and clearance to conduct the study was obtained from relevant authority before beginning the study. Since the total number of young infant admitted at the respective BMC premises (E4, neonatal Unit, C5, E5, C6, neonatal intensive care unit, and C2 wards) per month is small, therefore, a census sampling technique to ensure all eligible subjects are included during the study period was chosen. To supplement information on the young infants, a questionnaire-administered questionnaire was used to collect information from the infants’ mothers.

Data were managed using Microsoft Excel database, and were cleaned and imported to STATA version 13 for analysis. Multivariate logistic regression analysis done, significant levels was set at p-value less than 0.05.

**Results:** A total of 525 young infants were enrolled in the study from February to May 2021, of which 298 (56.8 percent) were male. NTDs was found in 73 (13.9%) of the 525 infants hospitalized at BMC during the study period, and the most common pattern was myelomeningocele 50(68.5%). Failure to use folic acid supplementation during preconception and early first trimester, living in rural residence, lower education and overweight during pre-conception period are the key factors associated with NTD among young infants

**Conclusion:** This study demonstrated that NTD is one of the prevalent condition contributing to admission among young infants at BMC.

**Recommendation:** BMC should provide regular health education on NTD prevention to all women of reproductive age in the community. The use of folic acid supplement in preconception should be emphasized to all women of reproductive age since folic acid use have been reported to reduce NTD in various studies including the present study.

## Background information

Neural tube defect (NTD) is a central nervous system (CNS) congenital malformation that occurs when the neural tube fails to close spontaneously between the 3rd and 4th week of uterine development(1). The physiological changes take place during the 3rd and 4th week of development and if not well developed they end up with the defect(2).NTD classification is based on embryological thoughts and the occurrence or absence of exposed neural tissue, as open or closed type. The condition presents in the form of anencephaly, encephalocele, or spinal bifida (myelomeningocele or meningocele). Environmental and genetic factors contributing to NTDs occurrence have been extensively explored in the previously studies. Furthermore, folic acid deficiency in pregnant women is reported to exaggerate risk of getting a newborn with NTD(3). It is estimated that more than 300,000 babies are born each year worldwide with NTDs, with a global prevalence of 18.6 cases per 100,000 live births(4). In Europe the prevalence was 9.1 per 10,000 life birth(5). The situation has been increasing overtime in African continent. Currently, the prevalence in African continent is approximated to be 50.74 per 10,000 birth(6), which is higher than that was reported in 2015 (7).The prevalence is even worse in sub-Saharan Africa(8) including East African countries, the prevalence is 84.84 cases per 10,000 birth (9). A studies done in Dar es Salaam, Tanzania, reported the prevalence of 30 per 10,000 live birth(10, 11).

Despite supplementation of vitamins and folic acid to pregnant women, most times are late to stop the development of the defect in the early pregnancy since women often delay their first prenatal visit (12). However, it is suggested that any woman of childbearing years take folic acid vitamins before becoming pregnant as the defects occur before most woman know they are pregnant and go to their first prenatal visit in late stages(13). Pregnant women should start antenatal visits early even before 12 weeks from conception and should continue to adhere to attending at least eight clinic visits (8). However, this remains ideal in the Northwestern Tanzania. This may be due to several factors such as being single and lack of education on early antenatal visits and poor social economic status(14, 15).Thus late reporting to the antenatal care could be the reason for increasing the preventable diseases like NTD. Therefore, education provision in the community is vital to improving the uptake of reproductive health care serves before conception, and thus reduce the incidence of preventable conditions including NTDs. However, in Africa, the high burden of NTD report show that about five per 1000 newborns have NTDs, and East Africa are most affected with nine per 1000 newborns(16). The rates of NTDs have shown to increase overtime in Africa(7, 17) that has been shown to causes high morbidity and mortality of neonates and infants during the first year of life(18). In the Lake Zone particularly at Bugando Medical Centre (BMC), cases of NTDs have been reported being among the common cause of morbidity and mortality (19). However, there is limited information on the prevalence, patterns and associated factors for NTDs in this setting. The information could be useful in developing interventions for improving quality care to reduce morbidity and mortality, and reduce the occurrence. Therefore, this study aimed to determine the prevalence, patterns and associated factors for NTDs among young infants admitted at Bugando Medical Centre. The information gathered in this study could help in knowing the burden of NTDs, setting priority planning for quality care to the affected younger infants and have to increase knowledge and awareness in setting strategies for prevention of NTDs in this setting as well as served catchment hospitals, who receives service at BMC.

## Material and methods

This was a hospital based cross sectional observational study conducted to determine the prevalence, patterns and factors associated with NTD among young infants admitted at Bugando Medical Centre in Northwestern Tanzania. The study was conducted from February to may 2021.This study included all admitted young infants aged 0-3months of life and they are respective mothers who provided consent in the study. Furthermore this study included all young infants with and without NTD who were referred from peripheral hospital to BMC. The dependent variable was NTD and independent variables were the associated factors including participant’s socio characteristics. Ethical clearance to conduct the study requested from Joint CUHAS/BMC Research Ethics and Review Committee (CREC) obtained with CREC/456/2021.Permission to conduct the study were requested from the director of Bugando Medical Centre. A written informed consent was requested from the mother after explaining the importance of the study before enrollment obtained. Any refusal to participate did not affect the services provided at BMC. Confidentiality was maintained throughout to the study. Mother of the young infants had the right to withdraw their infants at any time during the study.

## Data management and analysis

Data on each patient collected by an interviewer-administered questionnaire from mothers of the young infants admitted to Bugando Medical Centre. The questionnaire tool consisted of socio-demographic factors, clinical/obstetric factors as well as maternal awareness and knowledge of NTD. We conducted the interviews in their respective wards after the mother agreed to participate in the study. Training to four research assistants on the research tool was conducted. The training of research assistant included creating good rapport to participants, confidentiality of information gathered, correctness of the data collected and recoding client hospital registration number. The research assistants were allocated in the specific pediatric unit, E6, C5, neonatal unity and NICU. Principal investigator had to work together with research assistants and check the completeness of data collection tool.

## Validity and reliability of the study tool

The data collection tool was developed as the previous published studies (8, 20–22). This questionnaire tool was tested and refined to ensure validity and reliability of the tool used, and therefore the study findings.

## Results

### Young infants and their respective mother’s socio-demographic characteristics

As summarized in Table 1, this study involved 525 young infants and their respective mothers. Median age infants was six [IQR 2 – 60] days. Among the young infants, majority of them were male (298 (56.8%)), neonates (320 (60.9%)) and had normal birth weight (465 (88.6%)). The majority (n=55, 75.3%) the infant affected with NTD were from rural residence. Among the 73 infants with NTD admitted at BMC were from various regions in the Northwestern zone, including Mwanza 22 (30.1%), Kagera 12 (16.4%), Mara 6 (8.2%), Shinyanga 8 (11%), Geita 5 (6.8%), Simiyu 7 (9.6%), Katavi 3 (4.1%), Tabora 4 (5.5%), Kigoma 5 (6.8%) and Singida 1 (1.4%). Mwanza region had majority of NTD infants 22 (30.1%) followed by Kagera region 12(16.4%). Likewise, maternal demographic characteristics found that, 289 (56.8%) mothers were aged between 20-29 years. Most them were para three 147 (28.0%) and from urban 365 (69.5%). Moreover, majority of mothers whose their children had NTD were from rural residence (n= 55/73 (75.3%). Of all mothers 482 (89.9%) were married, and 272 (51.8% had not used folic acid before conception. Out of mothers identified to be HIV positive (17 (3.2%), 13 (76.5%) started ARV before conception of the current pregnancy.

**Table 1:** Socio-demographic characteristics of infants and their respective mothers (n=525)

| **Young infants Characteristics** | **Number (%)** |
| --- | --- |
| **Age of the newborn** |  |
| Neonates | 320 (61.9) |
| Infants | 205 (39.1) |
| **Sex of the newborn** |  |
| Male | 298 (56.8) |
| Female | 227 (43.2) |
| **Young infant birth weight** |  |
| Very low birth weight | 5 (0.9) |
| Low birth weight | 43 (8.2) |
| Normal birth weight | 465 (88.6) |
| High birth weight | 12 (2.3) |
| **Mother of the infant characteristics** |  |
| **Age of the mother** |  |
| <20 years | 6 (1.1) |
| 20-29 | 298 (56.80) |
| 30-39 | 211 (40.2) |
| 40-49 | 10 (1.9) |
| **Parity of the mother** |  |
| Para 1 | 90 (17.4) |
| Para 2 | 100 (19.1) |
| Para 3 | 147 (28.0) |
| Para 4 | 111 (21.1) |
| **Para 5 and above** | 77 (14.7) |
| **Place of residence** |  |
| **Urban** | 365 (69.5) |
| **Rural** | 160 (30.5) |
| **Marital status of the mother of the infant** |  |
| **Married** | 472 (89.9) |
| **Window** | 10 (1.9) |
| **Divorced** | 13 (2.5) |
| **Single** | 30 (5.7) |
| **Level of education of the mother** |  |
| **Did not go to school** | 44 (8.4) |
| **Incomplete primary school** | 35 (6.7) |
| **Complete primary school** | 177 (33.7) |
| **Incomplete secondary school** | 94 (17.9) |
| **Complete secondary school** | 110 (20.9) |
| **Diploma** | 37 (7.0) |
| **University** | 28 (5.3) |
| **Mother occupation** |  |
| **Farmer** | 158 (30.1) |
| **Housewife** | 128 (24.4) |
| **Employed** | 70 (13.3) |
| **Business** | 152 (28.9) |
| **Students** | 17(3.2)) |

**Figure 1.**
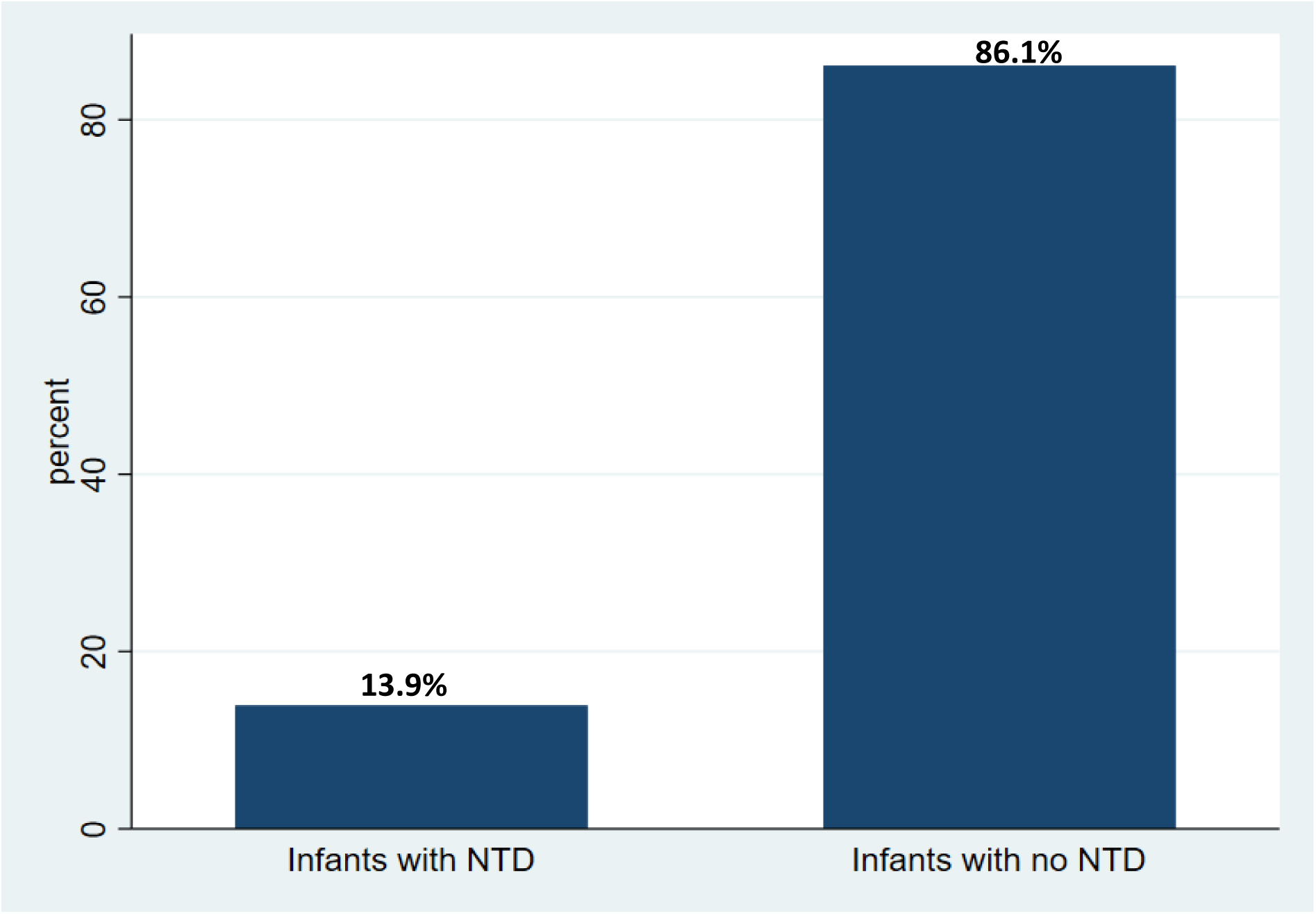
The prevalence of neural tube defect at Bugando Medical Centre.

**Figure 2.**
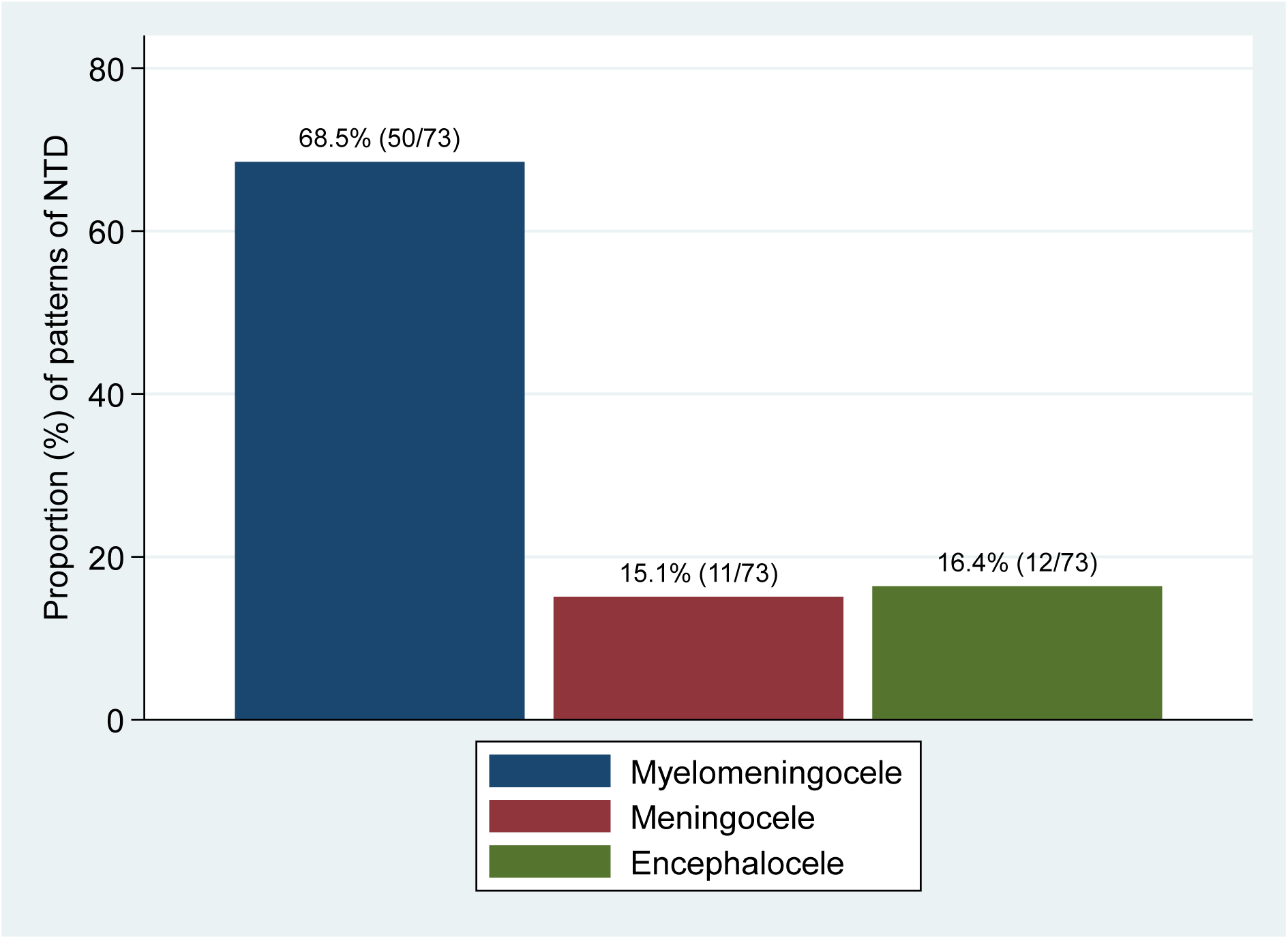
The pattern for Neural Tube Defect among Neural Tube defect infants admitted at Bugando Medical Centre during the study period.

### Prevalence of neural tube defect among young infants admitted at BMC

As summarized in Figure 4, among the 525 young infants admitted at BMC during the study period from February to May 2021, 73 (13.9%) infants were found to have NTDs.

### Patterns for Neural tube defect among admitted NTD infants during the study period

As summarized in Figure 5, the most observed pattern for NTD at BMC was myelomeningocele 50 (68.5%).

### Associated factors for neural tube defects among young infants admitted at BMC

As summarized in Table 2, univariate logistic regression analysis results show that the factors associated with NTD include Maternal age 20-29 years (OR= 0.1 [95% CI= 0.02-0.7]; p-value = 0.014), a high maternal parity of at least para 5 (OR =2.7; [95% CI = 1.8 – 6.7]; p-value = 0.003), living in rural residence (OR=10.1 [95% CI =5.68 – 17.9]; p-value < 0.001), primary education (OR =3.5 [95% CI =1.9 –6.4]; p-value <0.001, and informal education (OR=12.0 [95% CI =5.5 – 26.3] p-value <0.001). Other factors such as unplanned pregnancy (OR = 4.5; [95% CI = 2.7 – 7.6]; p-value <0.001), n^1^o^3.^h^9^i^%^story of folic acid use during first trimester (OR =2.1; [95% CI= 1.3 – 3.5]; p-value = 0.003, lack of information for the reasons of using folic acid before conception (OR= 3.3; [95% CI=1.9 – 5.8]; p-value <0.001) and mothers being overweight during pre-conception period (OR =2.4; [95% CI =1.4 – 3.9]; p-value < 0.001), were found associated with NTD.

**Table 2:**
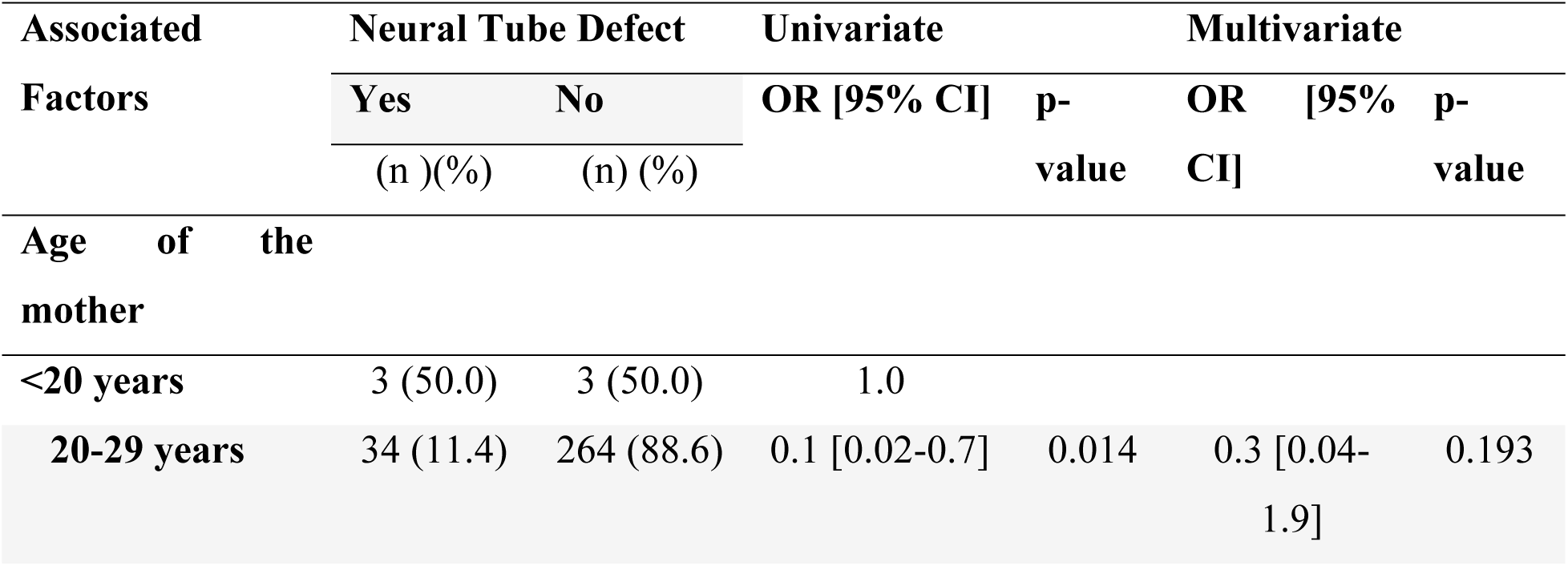
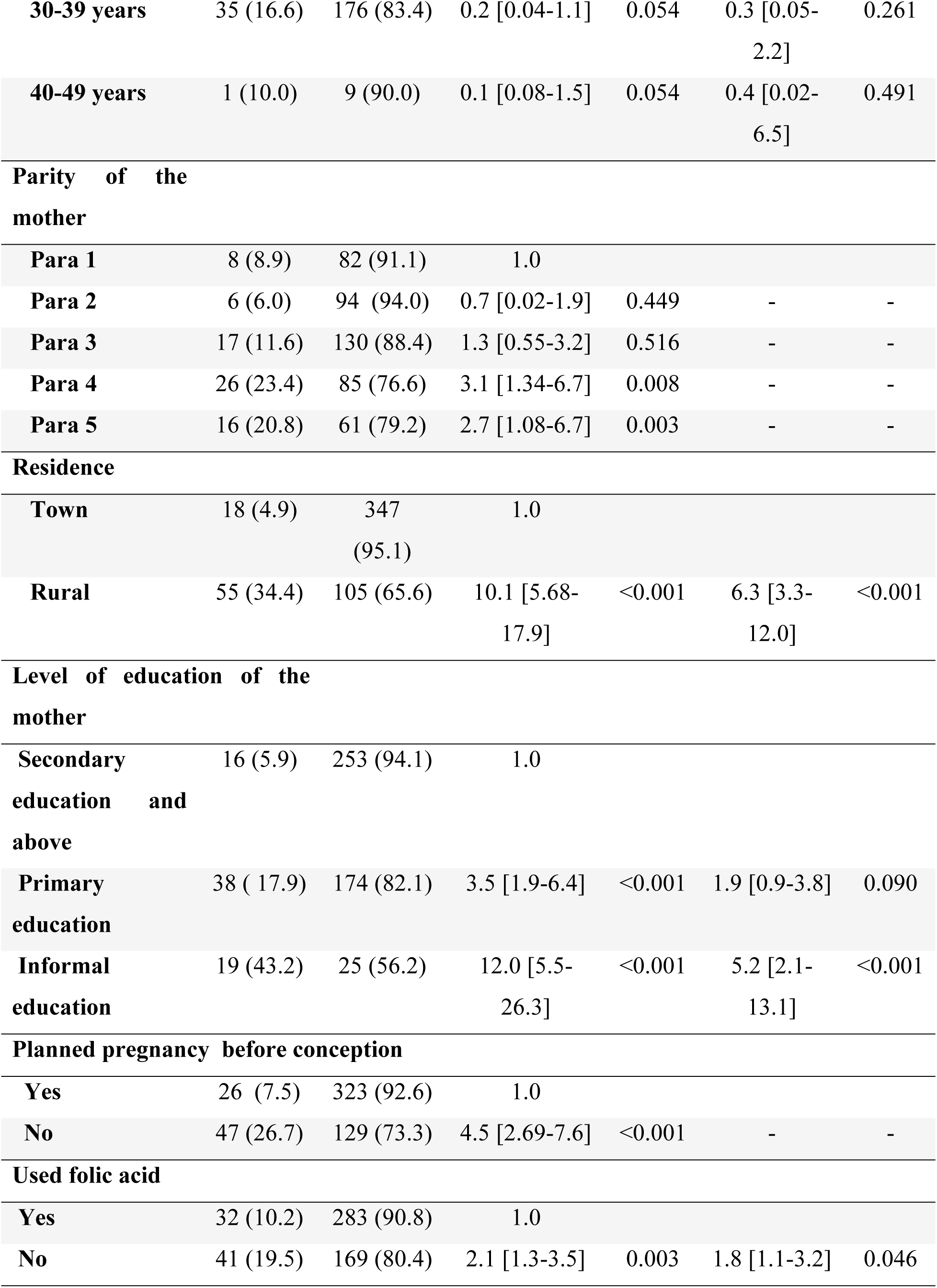
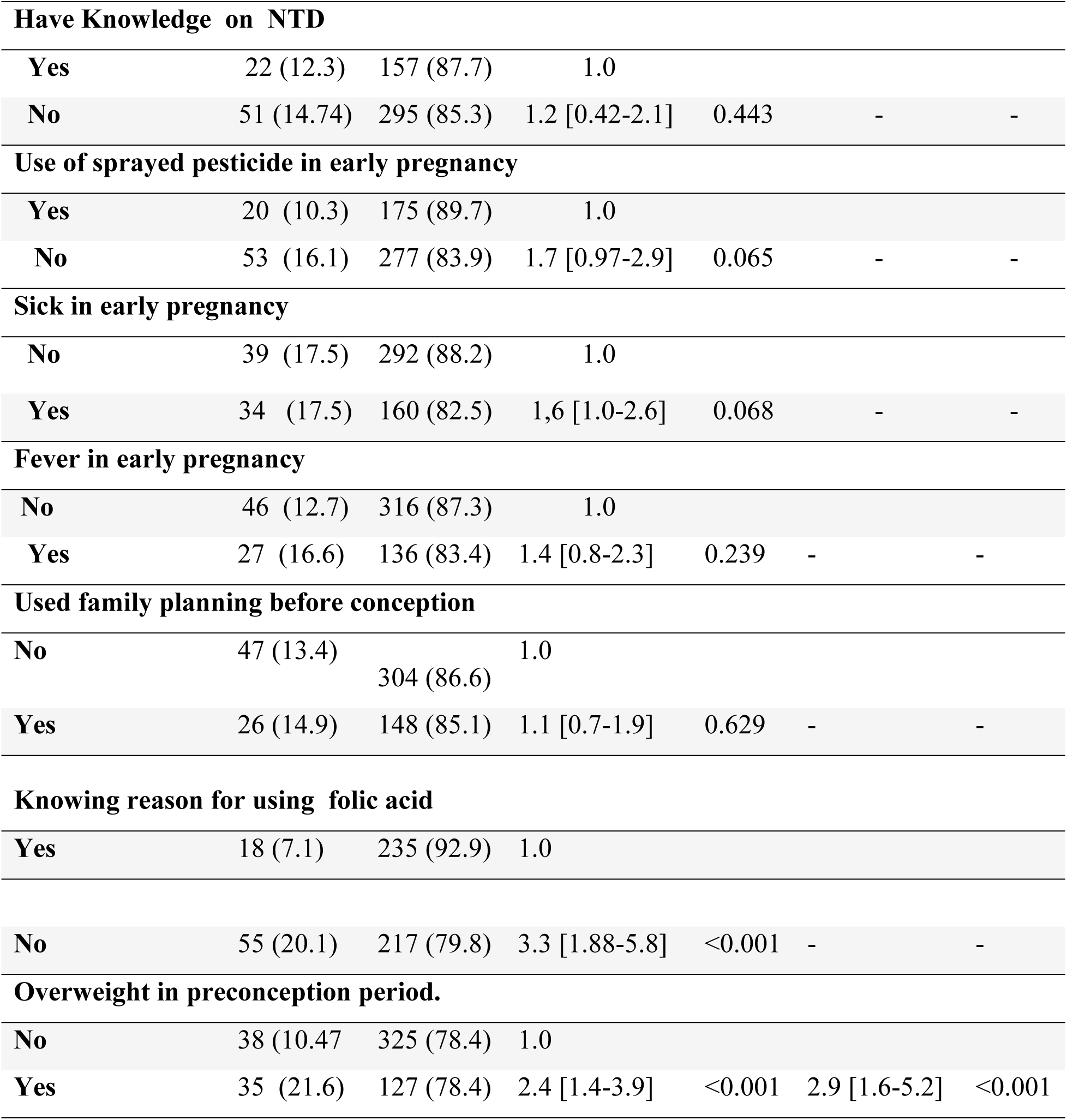
Associated factors for neural tube defect among young infants admitted at BMC (n=525)

Awareness on folic acid use, maternal parity, planned pregnancy and maternal age variables were eliminated in the final multivariate logistic regression model since the parity had collinearity with maternal age while planned for pregnancy and awareness on folic acid had collinearity with use of folic acid. In using multivariate logistic regression analysis, the factors found to be associated with NTD include not using folic acid supplement in the first trimester (OR= 1.8 [ 95% CI =1.1 – 3.2 p-value = 0.046), living in rural residence (OR=6.3 [95% CI = 3.3-12.0]; p-value <0.001), mothers with informal education (OR = 5.2 [ 95% CI= 2.1 – 13.1]; p-value < 0.001), and mothers being overweight during preconception period (OR = 2.9 [95% CI=1.6-5.2]; p-value < 0.001).

## Discussion

Our study found that neural tube defect is one of the most prevalent conditions contributing to admission of young infants at BMC. About patterns for NTD, myelomeningocele was the leading pattern at BMC. Moreover, living in rural residence, mothers with informal education, mothers who had being overweight prior conception and those without history of folic acid use in the first trimester were associated with increased risks for NTD.

### The prevalence of neural tube defects among young infants admitted at BMC

In our study, about 73 (13.9%) of all 525 young infants which is equivalent to 26.5 per 1000 live birth admitted at BMC during the study period were found to have NTDs. The prevalence reported in the present study is approximately similar with the findings of the previous study conducted by Gedefaw and Colleagues (8).

The prevalence reported in the present study is contrary to the findings of the previous studies from other areas (6, 23–26). BMC is the referral and consultant Hospital in Lake Zone, which performs specialized care as well as provision of pediatric surgical repair for NTD and other surgeries. Most previous studies have reported the prevalence of between 4.05 per 1000 total births and 63.3 per 10,000 live births (24, 27). Even, the previous study with slightly similar methodology to our study, on pattern and associated factors for NTD among young infants have reported high prevalence. The differences in prevalence of NTDs among young infants could be contributed by nutritional factors, environmental factors, and lack of routine use of folic acid supplement, lack of diet fortification program to enhance folic acid to mothers who delayed to start first antenatal visits. Mothers who received folic acid supplement at the clinic timely were protective of NTD (28). Different findings were reported by a study conducted in Kenya that found there were lower observed prevalence for spinal bifida and encephalocele whereby the study used different design and methodology. The study differed with our study as the study was conducted at AIC Kijabe hospital with bed capacity of 67, which was case a based hospital study that only patients seeking care to this hospital recruited in the study. Likewise, our study was a cross sectional observational study involving all infants admitted at BMC including referred cases. Therefore the finding is different from our findings as well as the previous study done in Kenya (29).

### Patterns for neural tube defects among young infant admitted at BMC

This study found that NTDs cases were 73 (13.9%), and male infants were most frequent affected 38 (52.1 %) than female infants 35 (47.9%). Our finding are similar to other studies who compared gender and development of NTD (19,30,31). Moreover, a cross sectional hospital based study conducted in Ethiopia, showed that male infants were at higher risks for NTDs than female infants were(32). Moreover myeleomeningocele was the most common type accounting for 50 (68.5%), followed by encephalocele 12 (16.4%) and meningocele 11 (15.1%). None of the infants had anencephaly in our study. This study finding are similar to other studies, where myeleomeningocele were found to be the leading NTD pattern. A study conducted in Adds Ababa reported myelomeningocele 91( 51.4%) followed by anencephaly 77 (43.5%)(24). An observational hospital study done in Sudan, reported that myeleomeningocele was the leading NTD 39 (42.9%), meningocele 35(38.5%), encephalocele 16 (17.6%) and anencephaly 1(1.1%)(33). Similarly, a study conducted at Tikuri Anbessa hospital in Ethiopia, found myeleomeningocele as the leading (64.4%) followed by meningocele (18.3%) and encephalocele (13.0%)(34). Furthermore, myelomeningocele (86.8%) was reported to be the leading type in the study conducted in Nigeria (35). Different findings were indicated by a study done in Tigray Northern Ethiopia found that anencephaly and spinal bifida were the leading 66.4 and 64.4 per 10000 births (32), respectively. The possible reasons of having increased cases of myelomeningocele could be type of nutrition taken by the mother, lack of awareness on the reason for using folic acid before conception, lack of education on the importance of folic acid supplements, geographical location as well as biological factors. One of the previous studies reported different findings from our finding that anencephaly 16 (23.5%) and meningoencephalocele 12 (17.7%) type of NTDs. The study mentioned the cause could be due to different environmental exposure, inadequate use of preconception folic acid supplementation and nutritional diet during the pre-conception period and social economic factors respectively (36). The other study conducted in Ethiopia, reported different findings as Anencephaly was the common type 68 cases per 10,000 births and spinal bifida 51 cases per 10,000 births and encephalocele 7 cases per 10,000 births which found to be common NTD type (8). This finding differs with our study finding as could be influenced by racial differences, geographical differences, nutritional factors biological genetics and social economic factors this study was done among births in the hospital.

### Associated factors for neural tube defects among younger infants admitted at BMC

In the present study, we found that not using folic supplement in the first trimester increased risks of NTDs. Almost 19.5% of mothers who reported not using folic acid supplement in the first trimester had infants with neural tube defect. Moreover, the study found that mothers who reported using folic acid before conception only 1(1.4%) had neural tube defect infant. The study highlighted that among 315 who used folic acid in their first trimester 32 (43.8%) of infants developed NTD. Of all, 210 not using folic acid in first trimester 41(56.2%) had NTD infants. About 214 (40.8%), mothers started their first antenatal at 24^th^ weeks of gestation. This means that most women delay in starting their first antenatal visit leading to inadequate prevention of NTD. The study found that most women who did not use folic acid in the first trimester had increased risks for NTD. Furthermore, these finding are similar to the previous studies done in developed countries. The study conducted in Cameroon, reported the similar findings that the lack of use of folic acid before conception and during early pregnancy is associated with increased risks of NTD. Furthermore study done in Nigeria, reported that not using folic acid supplement in first trimester have increased risks in development of NTD (1, 35). A systematic review and meta-analysis conducted in 2021 found similar findings that the lack of use of folic acid has increased risks for neural tube defect (17). Moreover, the study conducted in Mwanza on congenital anomalies, reported the similar findings that the proportional of mothers who used pre conception folic acid supplement in their first trimester was very low and were associated with congenital anomalies including NTD (37). However, these result findings could be associated with lack of knowledge on reasons for using folic acid, lack of awareness on food that supplement for folic acid, environmental factors, and delay in starting the first antenatal visit, failure to attend reproductive services before conception. Thus, in this study majority of mothers started the first visits at 24^th^ week of gestation. Likewise results highlights that timely use of folic acid prevents development of NTD. Additionally, a systematic review literature conducted by the previous study reported the use of fortified flour with folic acid supplement reduces neural tube defects (38). Moreover, others studies like that done in Sudan reported the use of folic acid had insignificant association with the development of neural tube defects (33). However, this study emphasized the importance of folic acid and/or multivitamin supplementation in preconception period and during the first three months of conception could help in prevention of NTD. These results are in agreement with other reported studies international findings (8, 23, 24, 37, 39, 40).

Furthermore, the study results found that the majority of mothers with young infants admitted at BMC 177 (33.7%) have primary education; mothers with university level were 28 (5.3%). Moreover, mothers with informal education found to have increased risk for neural tube infants than the educated women. In our study findings, the reason could be that, informal or less educated mothers have limited knowledge on important information for nutrition, inadequate knowledge in preventing the NTD, not knowing reasons for using preconception medication and limited social economic status. Likewise, a population based control study conducted in California found that mothers with higher education were less risks of neural tube defect compared to those with lower education who had high risks for NTD (41). The research finding have shown that, low maternal education was associated with an elevated risk of NTD in children. Results showed that risks for NTD varied by the education profile of the neighborhood, women who did not graduate from high school and lived in less educated, neighbor hoods showed two fold-elevated risk for NTD than high school educated woman who lived neighborhood (41). The study conducted in Colorado in 2002 found that low education status has association with NTD and mentioned that low maternal education status is an important predictor of having a child with NTD (23, 41–43). Moreover, in the current findings, this indicate that low maternal education status is an important predictor of having a child with NTD. Furthermore, previous studies showed that mothers who had low education have 1.7 fold-increased risk of delivering an infant with neural tube defect. Likewise, a study conducted in Sudan reported that education level of mother play an essential function in enhancing the level of knowledge of those mothers about neural tube defect to prevent reoccurrence. The study conducted in Italian found the low education level was associated with increased risk for neural NTD (33, 42).

Besides, this study found that mothers who reported being overweight in a preconception period showed increased risks for NTD. Majority of mother who reported being over-weight in preconception period showed association with increased risks for NTD. Furthermore, the study found that 35 (47.9 %) NTD infants were born by mothers who reported had over-weight in their preconception period. This could be due to lack of awareness to mothers on reduction of weight before conception, inadequate knowledge on nutritional diet, environmental factors and inadequate protective effect of folic acid as it is limited to the over-weight and obese, which need to be supplemented by an increased dose of folic acid to meet the demand. Similar findings are also reported by Rasmussen who reported that maternal overweight and obesity has an increased risk of an NTD-affected pregnancy (44). Likewise, a cross sectional study conducted by Fatima reported the mothers who were over-weight in the pre-conception period, they were more prone to have neural tube defect infants than the mother with normal weight (8, 45). The reason for these findings could be, NTD protective effect of folic acid is limited to the overweight mothers as well as obese mothers and they need higher doses of folic acid supplement to accomplish related serum level. Furthermore, others studies found that despite the use of preconception folic acid, also weight reduction or increased doses for folic acid supplement could protect this kind of mothers to NTD infants (8, 44, 46). Likewise, a meta-analysis study conducted by the other researcher showed the increased weight than normal in preconception periods puts the woman at risk for neural tube defect (47).

Additionally, our study findings show that mothers who reported living in rural resident have increased risks of neural tube defect. This could be due to social economic factors, education backgrounds, and poor infrastructures to reach the health services in preconception period, environmental pollution and lack of information on prevention of NTD. Again, majority of women living in rural resident started antenatal care when they were more than 12 weeks of pregnancy. Similar findings from previous studies reported that due to geographical differences and social economic status, as well as nutritional consumption of the mothers, increase the risk NTD (48, 49).

### Limitations of the study

Participants recall and self-reporting of associated factors, such as weight and medicines used in preconception period could have introduced recall bias. Furthermore, the study were conducted at the tertiary referral hospital in Lake Zone where the NTD are cared as the only hospital providing the care to young infants with NTD in the Northwestern part of Tanzania, this could have brought bias. These limitations restricted the generalizability of the study findings.

### Clinical implication of the study

This study has created awareness on the situation of NTD prevalence, patterns and associated factors among young infants admitted at BMC. This could help in decision making during planning for immediate care to prevented NTDs-related complications including reduction of mortality rates.

### Conclusion and recommendations

This study demonstrated that NTD is one of the prevalent condition contributing to admission among young infants at Bugando Medical Centre. Myelomeningocele was found to be the commonest NTD type at BMC. Failure to get folic acid supplementation during preconception and early first trimester, living in rural residence, lower education and overweight during pre-conception period are the key factors associated with NTD among young infants admitted at BMC. However, this study recommends that: Bugando Medical Center should provide regular health education on NTD prevention to all women of reproductive age in the community. Furthermore, the use of folic acid supplement in preconception should be emphasized to all women of reproductive age since folic acid use have been reported to reduce NTD in various studies including the present study. Likewise, further study to explore factors influencing uptake of folic acid among women of reproductive age during preconception period is recommended.

## Acknowledgement

The authors express their sincere appreciation to CUHAS/BMC administration and my research supervisors, for giving their approval and assistance in conducting this research. In addition, the authors is grateful to infants and respective mothers who participated willingly in this study. Furthermore, appreciation to the Hanze Foundation Groningen Netherland and Ministry of Health Community Development Gender, Elderly and Children Tanzania for their financial support which has led to the completion of dissertation.

## Author Contributions

CJ, HD, RL, and FM conceptualized the study, collected and analyzed data and wrote the manuscript. BK, ED, did analysis of the data and reviewed the manuscript. AT, CC, AH, KM, HJ, JK, AM, FA reviewed the manuscript and provided expert inputs in the manuscript.

## Funding

None

## Data availability statement

The datasets produced and analyzed during the current study are accessible from the corresponding author upon realistic request.

## Declarations

### Conflicts of interest

None

## Ethical approval and consent to participate

Ethical clearance for conducting the study was requested from the joint CUHAS/BMC Research Ethics and Review Committee (CREC) obtained with CREC/456/2021. Permission to conduct the study was requested from the director of Bugando Medical Centre. A written informed consent requested from mother after explaining the importance of the study was obtained before enrollment. Any refusal to participate did not affect the services provided at BMC. Confidentiality was maintained throughout the study. Mother of the infants had the right to withdraw their infants at any time during the study.

## Notes

### Competing Interest Statement

The authors have declared no competing interest.

### Funding Statement

The author(s) received no specific funding for this work.

